# A systematic review assessing the state of analytical validation for connected, mobile, sensor-based digital health technologies

**DOI:** 10.1101/2023.05.22.23290371

**Authors:** Yashoda Sharma, Katerina V. Djambazova, Celine D. Marquez, Kate Lyden, Jennifer C. Goldsack, Jessie P. Bakker

## Abstract

**Background:** Despite the recent proliferation of digital health technologies (DHTs), there is a lack of formal, industry-wide standards to evaluate the performance of the product’s algorithm in terms of its ability to measure, detect, or predict a clinical state. The advancement and successful use of DHTs in medicine requires that all stakeholders – clinicians, patients, payers, regulators, pharmaceutical companies, and the medical products industry – have a common understanding of what it means when a DHT has been analytically validated.

**Objective:** We conducted a systematic review to assess the state of the science on analytical validation for DHTs, using the criteria established by the V3 Framework and EVIDENCE checklist.

**Methods:** The systematic review was conducted according to the PRISMA guidelines. A multi-tier PubMed search identified studies published between April 15, 2020, and April 15, 2023, on analytical validation of DHTs; thereafter, each paper was assessed against the EVIDENCE checklist items specific to analytical validation. All studies were required to report quantitative data collected from a connected, mobile sensor product for the measurement, diagnosis, and/or treatment of a behavioral or physiological function, and compare the outcome measures to an established reference standard.

**Results:** Of the 1201 papers identified in the literature search, we identified 303 reporting the results of a DHT analytical validation study. The most prevalent therapeutic areas of focus were neurological (26%), cardiovascular (18%), and sleep conditions (17%). Health outcome categories most frequently captured by DHTs were gait (15%), heart rate/rhythm (15%), blood pressure and/or arterial stiffness (11%), sleep staging (10%), and mobility (9%). Only 208 papers (69%) reported all components of the EVIDENCE checklist focused on analytical validation, with the exception of software version and race/ethnicity, thereby meeting our definition of high-quality evidence reporting.

**Conclusion:** We are encouraged by the emerging literature evaluating whether outcome measures assessed by DHTs adequately reflect the physiological or behavioral parameter of interest; however, the quality of reporting is not yet sufficient to ensure the advancement of digital clinical measures that are fit-for-purpose for all members of a defined population and eliminate the need for redundant studies. We recommend that journals publishing analytical validation studies require the use of the EVIDENCE checklist as a reporting standard for these manuscripts.

## Introduction

Digital health technologies (DHTs) can be valuable tools in clinical research and across the care continuum. The degree to which they provide clinical value depends on the quality and completeness of the “evidence stack” that supports the DHT. This includes evidence that (1) the sensor-level data are accurate and precise (verification); (2) the outcome measures adequately reflect the physiological or behavioral parameter of interest for all members of a defined population (analytical validation; AV); and (3) the outcome measures are clinically-relevant for a particular population and context (clinical validation; CV). Lack of consistent and transparent reporting of evidence in these three categories has been a major obstacle in timely development of DHTs, contributes to waste of health ecosystem resources, and reduces access to high-quality DHTs. Through transparency of evidence generation, and ensuring fidelity to the appropriate evidentiary standards, we can expedite the advancement of digital medicine, delivering on its promise of enabling high-value care at a lower cost.

The V3 Framework describes the process of verification, AV, and CV for the evaluation of connected, mobile, sensor-based DHTs [1]. Subsequently, the EValuatIng connecteD sENsor teChnologiEs (EVIDENCE) checklist was developed to define appropriate reporting criteria for studies evaluating these products [2]. The overarching objective of the checklist is to establish consistent, high-quality reporting of studies and to serve a similar purpose to checklists applied to reporting clinical trials [3], assessments of diagnostic accuracy [4], and systematic reviews and meta-analyses [5]. High-quality reporting allows for results to be more comprehensively interpreted, compared across studies, tested for reproducibility, and enables the selection and privileging of DHT’s in the market that will deliver measurable value to patients.

Despite the widespread adoption of the V3 Framework and the EVIDENCE checklist since publication [6, 7], the quality of reporting for AV is still unclear. From our observations and anecdotally from community partners and participants, existing AV studies are often duplicated due to a lack of robust methodology, and/or lack of generalizability to all intended users of the DHT, and/or the study report is missing critical details. This is supported by a previously reported paucity of inclusive AV studies in the peer-reviewed literature [8] and the downstream documentation of racial bias in digital measurement products [9].

The Digital Health Measurement Collaborative Community (DATAcc) is a U.S. Food and Drug Administration (FDA) Center for Diagnostic and Radiological Health (CDRH) collaborative community [10, 11], hosted by the Digital Medicine Society. In this project, DATAcc set out to benchmark the state of the science by conducting a systematic review to identify peer-reviewed papers describing AV studies of connected, mobile, sensor-based DHTs and assess the quality of study reporting by applying the EVIDENCE checklist to the papers identified.

## Methods

### Literature Search

AV is defined as the process by which the performance of an algorithm converting DHT sensor-level data to physiological or behavioral outcomes is assessed against an appropriate reference standard amongst a study sample representative of the complete population of interest [1]. All AV studies involve human participants and clinical/medical outcome measures; thus, PubMed was selected as the most appropriate search engine. The search terms were designed with multiple layers separated by “and” or “not” Boolean operators; within layers, terms were separated by “or” (Multimedia Appendix 1). Layers 1 and 2 used Medical Subject Headings (MeSH) terms, requiring that papers be indexed as human studies (Layer 1) relevant to at least one aspect of digital medicine (Layer 2). Layers 3 and 4 included keywords targeting connected, mobile, sensor-based DHTs and the AV process, respectively. The concept of ‘validation’ or ‘agreement’ of an outcome against a reference standard are foundational to AV studies following V3, and thus these keywords were included in Layer 5. After a preliminary search using these five layers, Layers 6 and 7 were compiled in an effort to exclude irrelevant papers based on publication type (Layer 6), MeSH headings (Layer 7), and keywords (Layer 8). Finally, Layer 9 limited the search to the period from April 15, 2020, (the V3 Framework publication date) until April 15, 2023.

Digital medicine is a young field in which terminology and definitions are in active development. As such, Layers 3 and 4 were designed to be sensitive rather than specific, in an effort to capture the breadth of terminology in use today. We excluded keywords that describe specific sensors (e.g. accelerometry), non-specific product types (e.g. actigraphy), or brand names to avoid bias resulting from elevating papers with well-known tools while not capturing niche or novel digital products.

### Systematic Review

A multistep review of the identified studies was conducted, following the PRISMA 2020 (Preferred Reporting Items for Systematic Reviews and Meta-Analyses) guidelines [5]. Eligibility criteria followed the PICO (Population, Intervention, Comparison, Outcomes) framework [12], described in Table 1.

**Table 1:** Eligibility criteria applied to identified papers

| Exclusion criteria applied to identified papers following the PICO framework |  |
| --- | --- |
| Population | 1. Exclude studies that do not capture data from human participants. |
|  | 2. Exclude studies that are based on *in vitro* data. |
| Intervention | 3. Exclude studies that do not include a sensor-based DHT. Technology under assessment must be:<br>• Connected, meaning that there is a wired or wireless method to move data from the tool to the clinic/lab for analysis.<br>• Mobile, meaning that the tool is capable of collecting data remotely without oversight from trained personnel. Note that the study itself may have been completed in the clinic/lab setting.<br>• Sensor-based, meaning that the tool contains at least one sensor that samples a physical construct such as acceleration, temperature, pressure, light, or sound.<br>• Used for purposes of measurement, diagnosis, and/or treatment of a behavioral or physiological function related to a disease state or physiological condition. |
| Comparator/s | 4. Exclude studies that do not compare the output of the sensor-based DHT against an appropriate reference standard. |
| Outcome/s | 5. Exclude studies that do not report a quantitative assessment of algorithm performance. |
|  | 6. Exclude studies in which the sensor-based DHT output has undergone manual manipulation by participants or investigators. |
| Other | 7. Exclude studies that appear to meet the above criteria but are published as an abstract without a full-text paper. |

The V3 Framework and EVIDENCE checklist were developed specifically for DHTs that are mobile, connected, and sensor-based, described in the original papers as Biometric Monitoring Technologies (BioMeTs). We followed the four components as described by Goldsack et al. (2020) [1] by requiring that the tools be connected (we excluded tools that did not have a digital method of data transfer; for example, products that displayed data on a user interface only), mobile (we excluded tools that were not developed for use outside of the clinical/lab setting, even though the study itself may have been conducted in-lab), sensor-based (we excluded tools that did not contain at least one sensor), and used for purposes of measurement, diagnosis, and/or treatment of a behavioral or physiological function related to a disease state or physiological condition. We applied the latter criterion liberally; for example, an AV study focused on sports performance was considered in-scope if the digital outcomes could conceivably be applied to a disease state.

Sensor-based DHTs may be used for the development of novel digital endpoints for which no appropriate ‘gold standard’ exists. Similarly, there are many entrenched reference standards that are recognized as being sub-optimal. When considering the comparator, we again took a liberal approach by excluding studies only when no comparator was used or when the chosen comparator was clearly not an acceptable reference standard. For example, studies that performed validation of a digital sleep endpoint against a sleep diary instead of the ‘gold standard’ polysomnography were excluded.

Titles and abstracts were independently screened for eligibility by one of two trained investigators (YS, KD). A subset of 20% was randomly identified for auditing, which was screened by a third independent investigator (JPB). Cases of disagreement were resolved by consensus across all three investigators. During this process, papers with insufficient or ambiguous information in the title and abstract were marked as being preliminarily eligible. During the full-text review, all papers identified for potential exclusion were reviewed by an independent reviewer (JPB).

### Data Extraction

First, we extracted descriptive information from all papers meeting eligibility criteria, such as the country of origin, participant health status, and outcomes assessed. Categories for therapeutic area/s, health outcome/s, and technology type/s were applied based in part on lists developed in prior work [13].

The four technology type categories were as follows: environmental (a standalone DHT which does not come into direct bodily contact); wearable (a DHT worn on the body for some period of time); implantable (a DHT surgically inserted under the skin); and ingestible (a DHT that is swallowed). Smartphone- or tablet-based DHTs were categorized as environmental if they were task-based (such as finger-tapping or reaction time tests) or wearable if they were strapped to the body during data collection.

Next, we extracted data aligning to the EVIDENCE checklist, which identifies 24 required and 8 preferred reporting items for assessing the quality of studies that follow the V3 Framework [1, 2]. Of these, we extracted 13 items that are specifically applicable to AV (see Table 2). Our objective was to assess the reporting quality rather than the study results; as such, many data fields were extracted as “yes/no” depending on their presence or absence in the paper.

**Table 2:** Data extracted from all studies meeting eligibility

| EVIDENCE item | Item description | Data extracted |
| --- | --- | --- |
| Ethics and informed consent | Include a statement that Institutional Review Board (IRB) approval or Ethics Committee review of the study documentation was completed. Indicate whether a written consent was obtained from the study participants. | Reported: Y/N |
| Participants | Define the recruitment strategy and inclusion/exclusion criteria | Included healthy participants: Y/N<br>Therapeutic area category <sup>a</sup> |
| Sample Size | Indicate how the sample size was determined for the number of participants. | Sample size |
| Make and Model | State the make and model of the connected sensor technology used for data collection. | Reported: Y/N<br>Technology type category <sup>a</sup> |
| Sensor Characteristics | Describe the sensor modality(ies) used for data collection in as much detail as possible. | Sensor/s described in the paper |
| Form Factor and Wear Location | Describe the form factor (physical shape) and wear location (precise anatomic position of sensor) | Form factor described in the paper<br>Wear location described in the paper |
| Algorithm Description | Describe in as much detail as possible the algorithm used for data analysis in the study. | Reported: Y/N |
| Software Manufacturer and Version Number | State the version number and manufacturer of any software used for data collection. | Reported: Y/N |
| Outcome Assessed | Clearly identify the outcomes to be measured. | Digital outcome measure/s reported in the paper<br>Health outcome category <sup>a,b</sup> |
| Reference Standard | Describe the standard to which the performance of the connected sensor technology is being compared. | Reference standard reported in the paper |
| Statistical Analysis | Describe relevant statistical analyses. | Reported: Y/N |
| Participant Demographics | Define demographics collected for study participants, specifically age, race/ethnicity, and gender. | Age: adults/pediatrics |
|  |  | Age reported: Y/N |
|  |  | Sex/gender reported: Y/N |
|  |  | Race/ethnicity reported: Y/N |
| Numbers Analyzed/Findings | Describe the study's findings, including missing data. |  |
| <sup>a</sup> Therapeutic area, technology type, and health outcome categories were selected primarily from lists developed in prior work. <sup>b</sup> Outcome measures were categorized; for example, blood pressure, pulse transit time, pulse wave analysis, and pulse wave velocity were combined into 'Blood pressure and/or arterial stiffness'. |  |  |

Finally, the remaining eleven items listed as required in the EVIDENCE checklist were not extracted, as they are not specific to AV studies. These items were: structured abstract, study rationale; study objectives; data collection protocol; description of participant flow; description of adverse events; four items describing the various components of a discussion; and description of the funding source and competing interests.

## Results

### Literature Screening

The PubMed search retrieved 1,201 studies (Figure 1). During the title and abstract screening process, a subset of 20% of papers were reviewed by two independent investigators. The investigators agreed on the eligibility for 79% of these papers, while the remaining 21% required group consensus. Learnings from the consensus process were applied when screening the remaining 80% of titles and abstracts, and any that were ambiguous progressed to full-text review. All papers identified for exclusion during the full-text review were reviewed by an independent investigator with 100% agreement.

**Figure 1:**
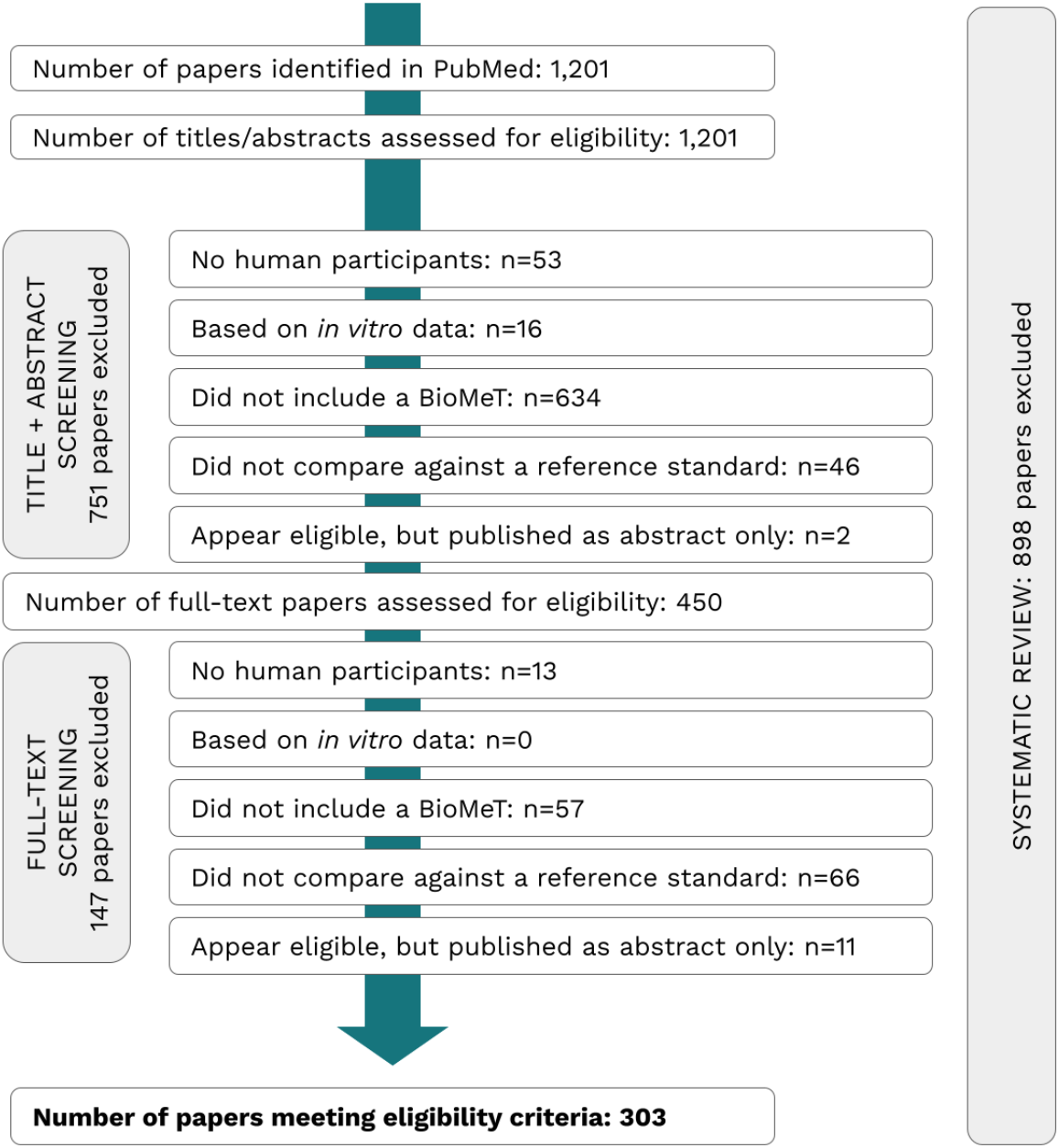
PRISMA Flowchart Identified studies were screened per PRISMA (Preferred Reporting Items for Systematic Reviews and Meta-Analyses) guidelines. In the first round of screening all papers were assessed for eligibility based on information contained in the title and abstract. A second round of assessment was conducted using the full-text of each paper.

Figure 1 describes the literature screening process, demonstrating that 898 papers (74.8%) did not meet PICO eligibility criteria. The remaining 303 papers progressed to data extraction.

### Descriptive data

Amongst the 303 eligible papers, we identified 38 unique therapeutic areas and 47 unique health outcome categories (Multimedia Appendix 3). The majority of studies 142/303 (47%) were categorized as ‘healthy or non-specific’ because they did not recruit participants on the basis of a disease condition. Of the 161 studies that recruited at least part of the sample on the basis of a disease condition, the most prevalent therapeutic areas represented were neurological (26%), cardiovascular (18%), and sleep conditions (17%).

The most common DHT technology types generating data for the 303 AV studies were wearables (69%), followed by environmental (27%), ingestible (2%), and implantable (1%); one study reported on both environmental and wearable DHTs. Amongst the wearable DHTs, form factors included straps or braces (reported in 130 studies), electrodes (41), cufs (39), watches (25), and adhesive patches (20). Categories identified for technology type, form factor, wear location, and sensor are presented in Multimedia Appendix 4.

Health outcome categories most frequently captured by these DHTs were gait (15%), heart rate/rhythm (15%), blood pressure and/or arterial stiffness (11%), sleep staging (10%), and mobility (9%). Note that health outcome categories were not mutually exclusive. Figure 2 displays the distribution of analytical validation studies across the most commonly studied therapeutic areas and health outcome categories, and the most commonly studied sensors for the top 10 health outcome categories.

**Figure 2:**
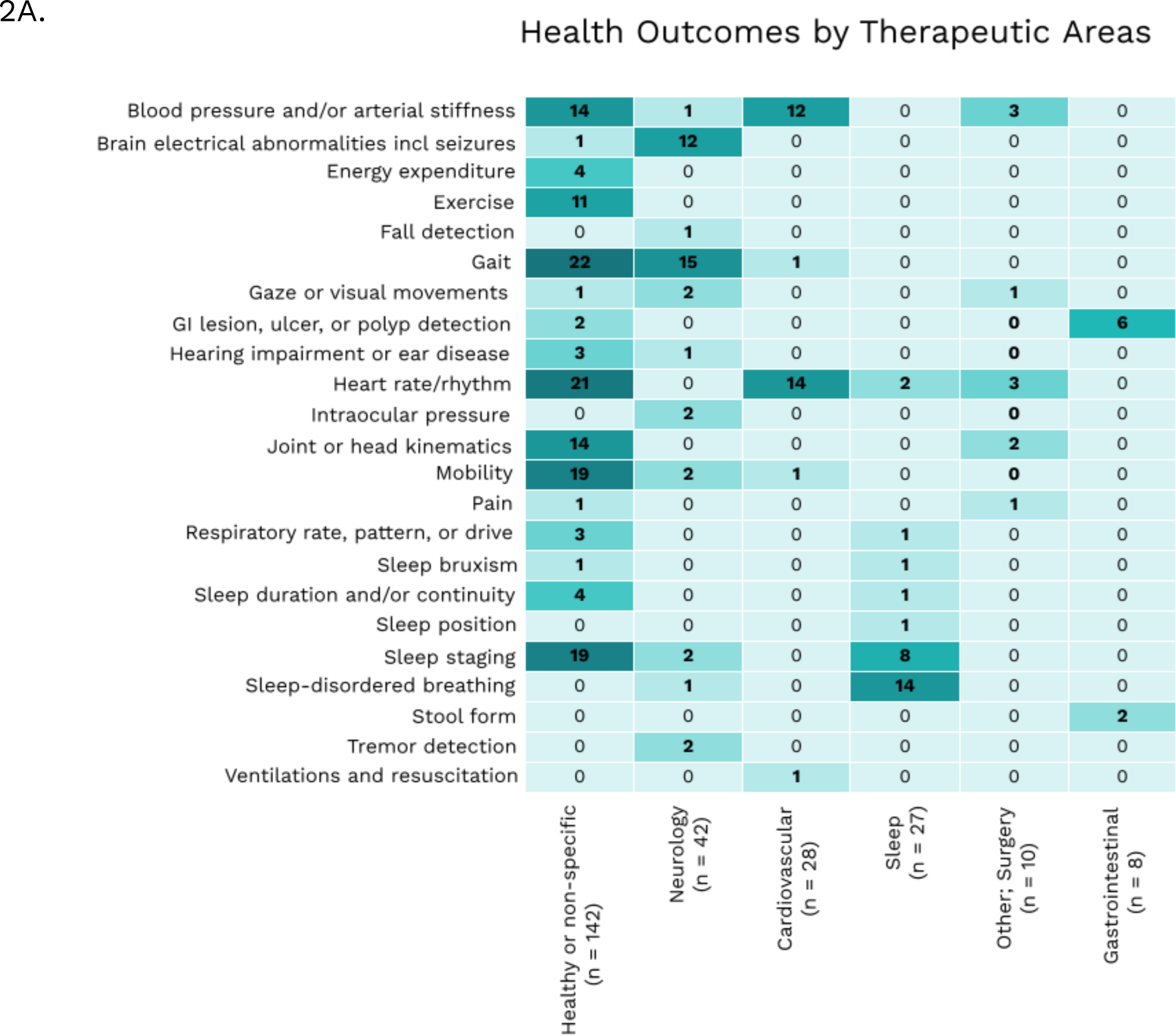
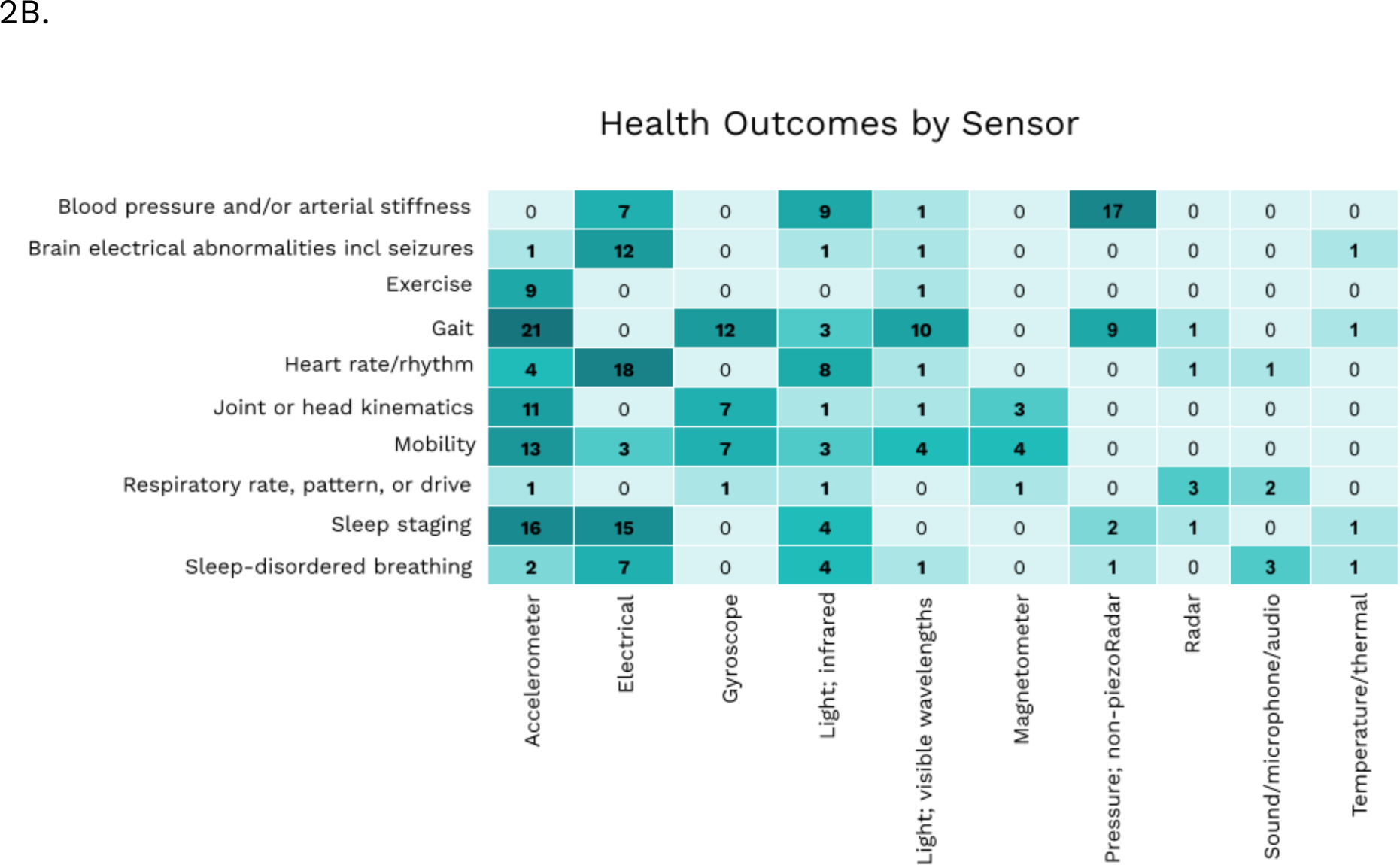
Distribution of Health Outcome Categories Studied by Therapeutic Area Distribution of analytical validation studies across the A) most commonly studied therapeutic areas and health outcome categories and B) most commonly studied sensors and health outcomes.

### Study reporting quality

By design, the eligibility criteria required that all papers focus on AV studies, which necessarily included a description of outcome measures and the statistical approach, and therefore, these EVIDENCE checklist items were complete for all 303 papers. Similarly, all papers reported a reference standard as this was part of the eligibility criteria (see subheading ‘systematic review’; above).

The number of papers reporting each remaining EVIDENCE checklist item are shown in Figure 3a. The number and percentage of papers not reporting the following items are as follows: ethics statement (4.3%); sample size (1.0%); DHT form factor and wear location (0.7%); algorithm description (17.8%); required demographic data (9%); and description of numbers analyzed including missing data (3.0%). In addition, the software version was not reported by 58.4%; however, not all DHTs have stand-alone software and so the extent to which this 58.4% represents missing information is not known. Participant demographics are a single EVIDENCE checklist item; however, we extracted each element separately and observed that 268 studies reported participant age (88%), 247 reported sex/gender (82%), and 27 reported race/ethnicity (9%); see Figure 3b.

**Figure 3:**
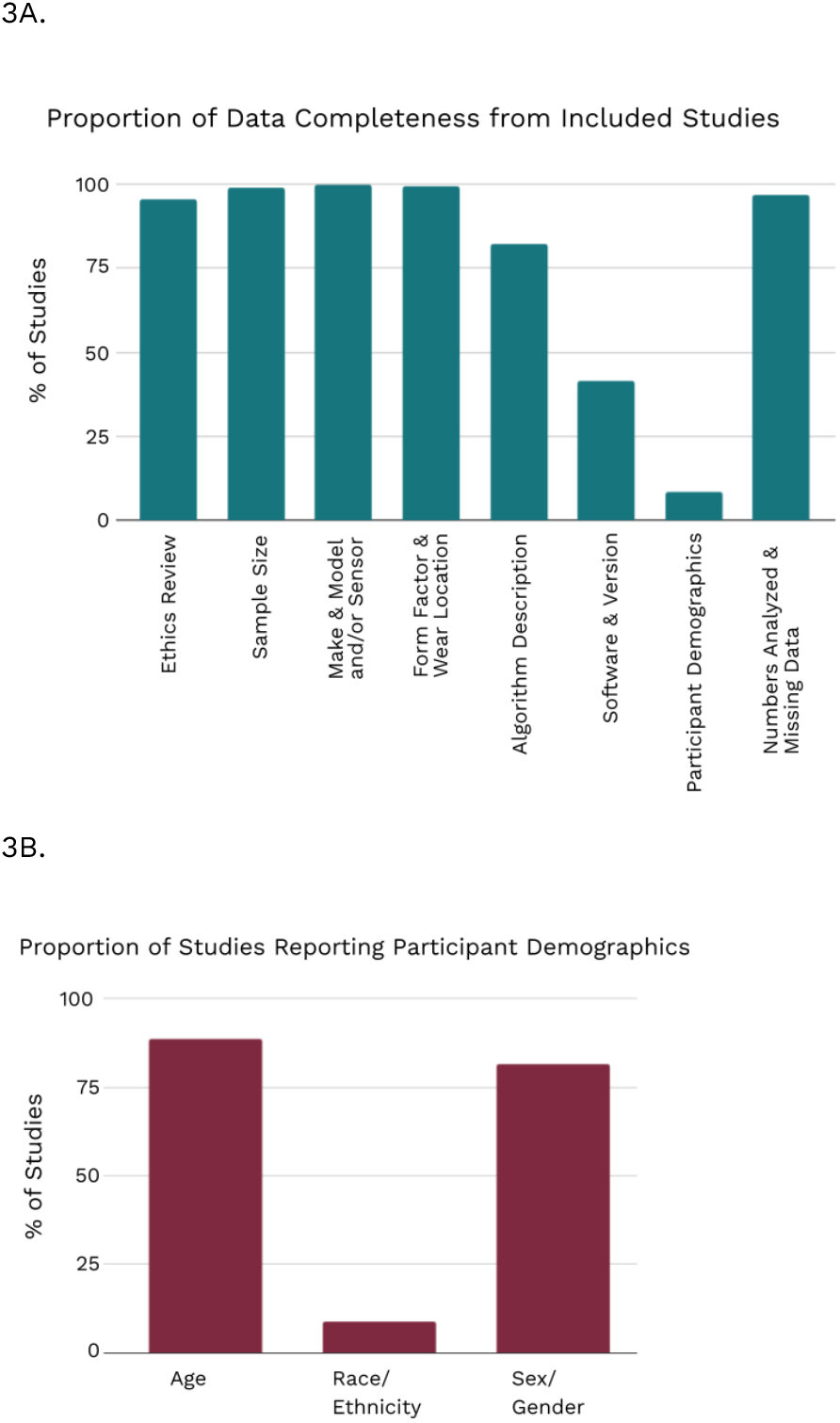
Assessment of Data Completeness Reporting completeness of the 303 eligible papers are presented (A) based on AV-specific EVIDENCE checklist items; and (B) based on individual elements of the participant demographics checklist item.

Those studies reporting all of the above EVIDENCE checklist items (Table 2), 208 papers (69%), with the exception of software version and race/ethnicity, met our definition of high-quality evidence reporting.

## Discussion

### Principal Findings

In the absence of industry standards or final regulatory guidance for analytical validation of connected, mobile, sensor-based DHTs, peer-reviewed studies can provide the evidence needed to ensure a digital product is fit-for-purpose. This systematic review was conducted to identify AV studies adhering to the V3 Framework and assess reporting quality of those studies per the EVIDENCE checklist.

In the three years since the publication of the V3 framework, over 300 studies were identified as adhering to the framework for analytical validation. However, the most notable trend that emerged during the literature screening process was that 46/498 (9.2%) of titles/abstracts describing a DHT, and 66/380 (17.4%) of full-text papers describing a DHT, were excluded due to the absence of an appropriate reference standard. Although some of the excluded studies focused on clinical rather than analytical validation, the majority did not capture an appropriate ‘ground truth’ for a given outcome measure. This included studies that reported inter-DHT agreement or back-validation studies in which an updated DHT or algorithm was compared against a previous model or version. As described in the Methods section, we took an inclusive approach to this exclusion criterion; for example, both automatic sphygmomanometry and invasive arterial pressure monitoring were considered appropriate references for blood pressure although the latter is widely considered to be the ‘gold standard’ [14]. Importantly, a reference standard based on self-reported data was not necessarily grounds for exclusion; for example, in some cases, pain intensity, stress level, and wear time may be best captured through self-report, in the absence of other known reference standards.

Although it is tempting to consider technology-based reference standards as ‘objective’, and therefore superior to self-report, many techniques such as polysomnography and electrocardiography continue to rely primarily on expert interpretation which presents challenges related to inter-rater variability [15, 16, 17]. Over time, some of the DHTs identified in this systematic review may themselves emerge as appropriate reference standards. This will first require a corpus of high-quality AV studies demonstrating consistently excellent results across generalizable samples and contexts of use. Recognizing that reference standard selection may not be straightforward during the development and assessment of novel outcome measures, we encourage investigators to select the best representation of ‘ground truth’ and include descriptions of the strengths and limitations of the selected reference standard. Future research is required to explore the selection of appropriate reference measures in the growing field of sensor-based DHTs to measure clinical outcomes.

Substantial variability in reporting completeness of the AV-related EVIDENCE checklist items was also observed (Figure 3). Most strikingly, of the 303 studies meeting PICO eligibility criteria, 277/303 (91%) did not report race/ethnicity. Seventy-eight percent of these studies were conducted outside of the United States, and we recognize that race/ethnicity reporting may not be mandated by funders or regulatory bodies in some regions. We observed that in place of race/ethnicity, some studies included Fitzpatrick Skin Phototype information [18]. While the Fitzpatrick scale has limitations, it is a widely used method for numerically classifying human skin color [7]. Several studies have demonstrated that wearable sensors, particularly those based on infrared light absorption, are not as accurate for people with varied skin pigmentation [19]. Therefore, it is important to recognize that race/ethnicity categories are often used as surrogates for other factors, such as skin tone, and in isolation these categories are not sufficient for assessing DHT generalizability. Similarly, although not required in the EVIDENCE checklist, we encourage authors to report additional descriptive information such as socioeconomic variables, language, and geographic disbursement that may be helpful for assessing generalizability.

While 53/303 (17%) studies did not report the DHT make/model, only 1% of studies failed to report either make/model or a description of the sensors; however, we observed substantial variability in the depth of information provided. The most helpful studies described the sensors in a way that allowed the reader to understand the underlying physical construct - such as pressure, sound, or temperature - along with the units and sampling frequency. Some papers described the use of an inertial measurement unit (IMU) which typically refers to the combination of an accelerometer and gyroscope with or without a magnetometer; as such, IMU was not considered an adequate description. Many identified studies reported AV of an algorithm that was not paired to a particular form factor or sensor; for example, an algorithm requiring heart rate that may be collected by any verified sensor using techniques such as electrocardiography, photoplethysmography, or pulse applanation tonometry. Several studies included a description of methodology rather than sensor/s; for example, pulse oximetry is a methodology but use of an infrared light sensor can be inferred. Finally, we observed that 58% of studies did not report the name and version number of DHT software; however, we were not able to diferentiate when this was a missing item versus not applicable for DHTs that do not include a stand-alone software package.

Overall, while it is encouraging to see the substantive emerging literature evaluating whether outcome measures assessed by DHTs adequately reflect the physiological or behavioral parameter of interest, the quality of reporting in the peer-reviewed literature is not yet sufficient to ensure the advancement of digital clinical measures that are fit-for-purpose for all members of a defined population and eliminate the need for redundant studies. With 112/1201 papers reviewed excluded due to the absence of a reference standard or inappropriate selection of a reference standard, the fundamentals of AV study design are lacking in the published literature as this information is critical to replicability. Similarly, more attention must be paid to inclusive study design and reporting given that 91% of AV studies did not report on race/ethnicity and 10% lacked any information on race/ethnicity, age or gender. This is unacceptable given the scientific rationale for doing so if devices are intended to be validated for use in a real-world population.

The EVIDENCE checklist is a comprehensive tool to support researchers, peer-reviewers, and editorial staf in improving the quality of published AV studies, similar to the improvements in clinical trial reporting driven by the CONSORT statement [20]. This approach is fundamental to ensuring that the rapidly-growing use of DHTs delivers on the promise of improved clinical decision-making and benefits all members of target populations.

### Strengths and Limitations

To our knowledge this is the first systematic review focused specifically on benchmarking the state of science of DHT AV studies. The sensitive nature of our search terms gives us confidence that most relevant papers were identified, although it is possible that some were missed due to highly heterogeneous terminology. Our assessment of study reporting quality was based on a peer-reviewed checklist, assembled by experts with cross-disciplinary expertise.

Alongside these strengths, several limitations are acknowledged. In attempting to align this review with the V3 Framework and EVIDENCE checklist, we excluded studies on DHT AV prior to 2020. We also limited our search to a single database focused primarily on the clinical literature; however, we were focused on studies of human participants in the medical field, and PubMed does include many engineering journals including >200 from the IEEE family. The sensitivity of our search terms resulted in the identification of >1,000 papers, and it was not feasible for independent reviewers to screen the entire list. Instead, we adopted a quality control process described elsewhere [12], in which a subset of titles/abstracts was audited for quality control. There was disagreement for approximately 20% of the titles/abstracts within this subset, which was largely due to lack of information within the abstract. As such, reviewers were instructed to err on the side of caution and allow any unclear abstracts to progress to full-text review. This process meant that papers excluded during title/abstract review were clear-cut, and all 147 papers excluded during the more thorough full-text review were verified by at least two investigators.

### Conclusions and Next Steps

Industry standards and regulatory guidance for AV of DHTs have not kept pace with the increasing use of digital clinical measures to inform clinical decision-making in healthcare delivery, clinical research, and individual health promotion. While the proliferation of scientific studies evaluating the processing of sensor data into clinically interpretable information is encouraging, the quality of reporting is inconsistent and inadequate. To ensure that DHTs generate data and information that are trustworthy, equitable, and fit-for-purpose for clinical decision-making, more value must be placed on the reporting of demographic information such as the definition of race and ethnicity in these studies and the science behind the selection and reporting of appropriate reference measures must be advanced. We recommend that journals publishing AV studies require the use of the EVIDENCE checklist as a reporting standard for these manuscripts.

In keeping with the mission of the Digital Health Measurement Collaborative Community [10], the 208 high-quality studies will be used to develop a searchable library. With the Analytical Validation Library, we will share studies that exemplify the quality of AV needed to advance digital health and ensure that digital health measurement products are fit-for-purpose. Future iterations of this work, including capturing studies prior to 2020, can inform the quality of studies in the library. As it grows, the AV Library will contribute to advancing the field by providing confidence in the use of these digital measurement products to capture accurate and reproducible clinical data for superior decision making to improve lives.

## Data Availability

Data are available upon request.

## Acknowledgments

The authors would like to thank the members of the Digital Health Measurement Collaborative Committee (DATAcc) Steering Committee and Analytical Validation project team for critical insights on study design for the systematic review and development of the Analytical Validation Library.

## Author Contributions

JCG and YS conceived the study. JCG, KL, CDM, and YS participated in study design. JPB, JG, KD and YS drafted the search terms and inclusion criteria. JPB and KD ran the search on PubMed. JPB, KD and YS participated in the screening and data extraction phases, and standardized the extracted data. JPB and YS carried out the data analysis and created visual representations. JPB, KD and YS drafted the first iteration of the manuscript. All authors added substantial details to the final manuscript, and read and approved the final manuscript.

## Data Availability

Data are available upon request.

## Abbreviations

AV: Analytical Validation
BioMeTs: Biometric Monitoring Technologies
CONSORT: Consolidated Standards of Reporting Trials
DATAcc: Digital Health Measurement Collaborative Community
EVIDENCE: EValuatIng connecteD sENsor teChnologiEs
IEEE: Institute of Electrical and Electronics Engineers
PRISMA: Preferred Reporting Items for Systematic Review and Meta-Analysis
V3: Verification, analytical validation and clinical validation

